# Urinary Cortisol and Cardiovascular Events in Women vs. Men: The Multi-Ethnic Study of Atherosclerosis

**DOI:** 10.1101/2023.07.21.23293023

**Authors:** Spencer Flynn, Preethi Srikanthan, Keeley Ravellette, Kosuke Inoue, Karol Watson, Tamara Horwich

**Author notes:** **Corresponding author:** Spencer Flynn, 10833 Le Conte Ave, Los Angeles, CA 90095.

## Abstract

**Background:** Research suggests that women experience greater cardiovascular ischemic effects from stress than men. Visceral adiposity is an endocrine tissue that differs by sex and interacts with stress hormones. We hypothesized that urinary cortisol would be associated with increased cardiovascular events and change in coronary artery calcium score (CAC) in women, and these relationships would vary by central obesity.

**Methods:** In the Multi-Ethnic Study of Atherosclerosis Stress Ancillary study, cortisol was quantified by 12-hour overnight urine collection. Central obesity was estimated by waist-hip ratio (WHR). Multivariable Cox models estimated the relationship between cortisol and cardiovascular events and assessed for moderation by WHR. The relationship between cortisol and change in CAC Agatston score was assessed by Tobit regression models.

**Results:** 918 patients were analyzed with median follow up of 11 years. There was no association between urinary cortisol and cardiovascular events in the cohort. However, in individuals with below median WHR, higher urinary cortisol levels (upper tertile) were associated with higher cardiovascular event rates in the full cohort, women, and men, but not in groups with above median WHR. There was significant moderation by WHR in women, but not men, whereby the association between elevated cortisol and increased cardiovascular events diminished as WHR increased. Urinary cortisol was associated with increased change in CAC in women (P=0.003) but not men, without moderation by WHR.

**Conclusions:** Our study highlights associations between cortisol and subclinical atherosclerosis in women, and moderation of the relationship between cortisol and cardiovascular events by central obesity in both genders.

## Introduction

Stress has been linked to cardiovascular disease (CVD) in epidemiologic, behavioral, interventional, and mechanistic studies.^1–4^ Stress results in the activation of the hypothalamic-pituitary-adrenal axis and the sympathetic nervous system, raising the levels of catecholamines and glucocorticoids, the most physiologically important of which is cortisol.^5,6^ Various mechanisms have been proposed for the role of cortisol in the development of CVD, including its contributions to hypertension, insulin resistance, hemodynamic stress on arterial walls, inflammation, atherosclerosis, and thrombosis.^7,8^ Potential behavioral sequelae of stress – low physical activity, high-fat diet, and smoking – likely also contribute to disease.^4^ Previous studies have found associations between increased stress – as measured by urinary catecholamines and cortisol– and plaque burden in coronary vessels, incident hypertension, and adverse cardiovascular events.^9–11^

Women and men may have different cardiovascular responses to stress. Sex differences in CVD risk have become more widely recognized, with stress emerging as a particular predictor of CVD risk in women. Studies have demonstrated that women with CAD are more susceptible to mental stress-induced myocardial ischemia (MSIMI), predisposing them to a greater risk of mortality and additional adverse CV events.^12,13^ Women have also been shown to have higher platelet reactivity to stress compared to men.^14^ Furthermore, postmenopausal women in particular may exhibit heightened hemodynamic responses to stress, such as elevated blood pressure and heart rate.^15–17^ This change following menopause may be attributable in part to the atherogenic lipid profile and increased visceral adiposity seen in post-menopausal women.^18^

Further modifying the relationship between stress hormones and cardiovascular disease is visceral adipose tissue (VAT). Visceral obesity is strongly associated with adverse cardiovascular outcomes and a broad number of cardiometabolic risk factors.^19,20^ Hypercortisolism is also associated with visceral obesity.^21,22^ This association is most apparent in disease states of pathologically elevated cortisol as occurs in Cushing’s syndrome, but it has also been shown that patients with higher VAT have higher 24-hour cortisol levels.^23–25^ VAT has been linked to an up-regulated hypothalamic-pituitary-adrenal axis (HPA).^26–28^ Women with increased visceral obesity have been shown to have resistance to low oral dexamethasone suppression test,^29^ and to have increased urinary cortisol.^30–33^ These studies highlight stress hormone resistance and upregulation in obese individuals.

In this study, using longitudinal data from the Multi-Ethnic Study of Atherosclerosis (MESA) Stress Ancillary Study, we aimed to investigate sexual dimorphism in the relationship of biological markers of stress and both subclinical atherosclerosis (as measured by change in coronary artery calcium (CAC) Agatston score) and cardiovascular events. Furthermore, we examined if these relationships varied by central adiposity as measured by WHR.

## Methods

MESA is a prospective epidemiologic study including 6,814 patients without cardiovascular disease at time of enrollment that began in 2000. Methods have been previously described.^11,34^ Briefly, its goal was to investigate the prevalence, progression, and risk factors for subclinical CVD in a large, multi-ethnic, population-based sample throughout the United States.^34^ The MESA Stress 1 Study was an ancillary study of 1,002 randomly selected white, black, and Hispanic patients from the New York and Los Angeles sites organized in conjunction with MESA examinations 3 and 4 (March 2004 to September 2005 and September 2005 to May 2007, respectively).^35^ Of these 1,002 patients, 918 patients had complete demographic and clinical data and were included in the present study. Both MESA and the Stress Ancillary Study were approved by the institutional review boards at the participating institutions, and all participants gave written informed consent.

### Quantifying urinary cortisol

The MESA Stress 1 collection process for urine samples and quantification of urinary cortisol has been previously described.^11,35^ Briefly, participants were instructed as to how to collect a 12- hour overnight urine sample. Following a previously used protocol,^36^ staff calculated the start time of the collection based on calculating backward 12 hours from the participant’s usual waking time. The participant was given detailed instructions to void at the start time and collect all urine until the stop time. Participants transferred each voided urine sample to a collection bottle containing a preservative (sodium metabisulfite; NA2S205, crystal form; reagent ACS grade, Fisher Scientific no. S244). Participants stored the collection bottle in a cool place and returned it to the study site as soon as possible. After collection, aliquots of urine were acidified with (∼6 N) HCl to a pH of <3 and frozen at -80 C until assayed.

Samples were analyzed for cortisol at Northwestern University using the urinary cortisol EIA kit. Intra-and interassay coefficients of variation were 3.2% and 9.99% for urinary cortisol. Urinary cortisol levels were all normalized by creatinine to reduce variability due to urine concentration.

### Categorizing clinical outcomes

Methods for assessing cardiovascular events and measuring CAC in MESA have been previously reported.^11,34,37,38^ New onset cardiovascular events were the primary endpoint and were identified beginning from a patient’s enrollment visit in the MESA stress study. Any patient with a cardiovascular event prior to their MESA Stress enrollment was excluded. A telephone interviewer contacted each participant (or representative) every six-nine months to inquire about all interim hospital admissions, outpatient diagnoses, and deaths. Two physicians reviewed all medical records for independent end-point classification and assignment of event dates. New onset cardiovascular events included all CVD and heart failure (HF) events adjudicated as part of MESA. Criteria for CVD included myocardial infarction, resuscitated cardiac arrest, definite or probable angina, stroke, stroke death, coronary heart disease death, atherosclerotic death, and CVD death.

CAC scans were performed on a randomly selected subset of one-half of the participants (n=447) by chest computed tomography (CT).^34^ Images were calibrated using a calcium phantom to control for variability between scanners, and the phantom was included in all images with parameters set to reduce differences in the signal-to-noise ratio.^39–41^ All CT scans were read at the Harbor-UCLA Research and Education Institute in Torrance, California. Coronary calcification was identified, quantified, and calibrated according to the readings of the calcium phantom. Each patient underwent 2 CT scans at each timepoint and the Agatston score was calculated for each scan (phantom adjusted) and the mean of these 2 CAC scores was used in the analysis. Intra-and inter-observer agreement was very high (kappa statistics 0.93 and 0.90, respectively).^39^ If there was a CAC measured at the same time point as the urinary cortisol measurement, then this CAC was used. If not, then a CAC measured at the closest time point within the MESA Stress examinations was used. The follow-up CAC was taken at MESA exam 5 for all patients. Change in CAC was determined as this later timepoint CAC minus the baseline CAC and was normalized to a per-year change.

### Body composition measurements

Anthropometric body composition measurements in MESA have also been previously described.^42^ Briefly, all participants were measured wearing light clothing and no shoes. Height was measured using Accu-Hite® stadiometers, rounded to the nearest mm. Body weight was measured using a Detecto Platform Balance Scale, rounded to the nearest pound. Waist and hip girth measurements were made in the standing position using Gulick II 150 cm anthropometric tape measures keeping the tape within a horizontal plane. No measurement was taken around clothing. Waist circumference was measured at the level of the umbilicus. Hip circumference was measured around the widest circumference of the buttocks. Waist-Hip ratio (WHR) was calculated from waist and hip girth measurements and used as our index of central obesity.^43^

### Statistical methods

All continuous comparisons between two groups used Student’s t-test, while categorical comparisons used chi-square tests. Cox regression analyses were performed to determine the relationship between urinary cortisol and new-onset cardiovascular events. All urinary cortisol levels were normalized to creatinine and then log base 2 transformed, so the estimated HRs represent an increase per doubling of cortisol. To assess the moderation by WHR on urinary cortisol, a multiplicative interaction term was included in all multivariable models. All WHR values were multiplied by 10 to improve model interpretability, and centered around the mean in order to conform to model assumptions. Cortisol was analyzed as categorical tertiles and as a continuous variable.

For the analysis of change in CAC by urinary cortisol, Tobit regression was used due to model overdispersion with a high number of 0 values. Change in CAC values were standardized to reflect annual change, and then ln(change in CAC/year + 1) transformed to conform to model assumptions. Outputs of the CAC models were annualized relative change values. Missing data was not imputed. Additional covariates were selected for the analysis of the relationship between urinary cortisol and new-onset cardiovascular events, as well as between urinary cortisol and change in CAC. These covariates were selected a priori and include age, race, annual income, smoking, alcohol, physical activity, systolic blood pressure, BMI, and low-density lipoprotein levels (see **Table 1** for further details on categories of covariates). All models included these covariates. Formal second-order moderation testing was also performed with gender and central obesity as moderators for the associations between cortisol and both CVD and change in CAC. All analyses were adjusted for multiple comparisons using the Benjamini-Hochberg method to control the false discovery rate. All statistical analyses were performed with R version 4.1.0. (R Foundation for Statistical Computing, Vienna, Austria).

**Table 1:**
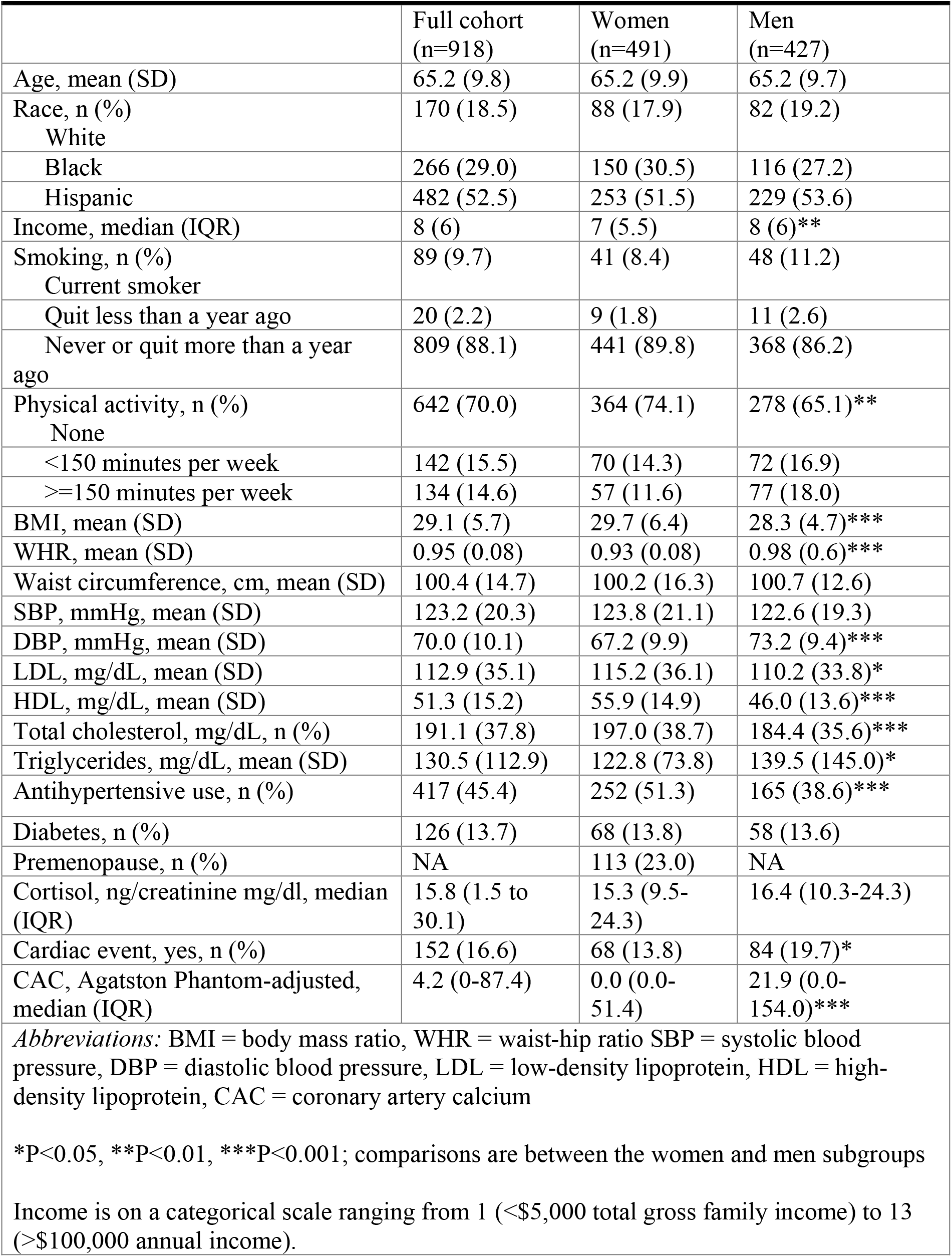
Baseline Demographics and Clinical Characteristics.

## Results

There were 918 total patients with complete demographic and clinical data without any CV events prior to MESA Stress enrollment, consisting of 491 women and 427 men (**Table 1**). The cohort was 52.5% Hispanic, 29.0% black, and 18.5% white. Mean BMI was 29.1 kg/m^2^, and BMI was significantly higher in women than men (29.7 v. 28.3, P<0.001). Mean WHR was 0.95, and was significantly lower in women compared to men (0.92 v. 0.98, P<0.001).

With regards to urinary cortisol, there was no significant difference in cortisol between men and women (**Table 1**). There were 152 patients (16.6%) who experienced new-onset cardiovascular events over the course of the study follow-up. Average follow-up was 11.02 (SD 2.99 years), with 5.46 (SD 3.30) years average time to new-onset cardiovascular events. Cardiovascular events were more common in men than women (unadjusted incidence percentages 18.7% v. 13.8%, incidence rate per 100 person years were 1.96 and 1.30, respectively, both comparisons P<0.05). Median CAC at baseline was also higher in men (21.9, IQR 0-51.4) than women (0, IQR 0-51.4) (P<0.001).

Patients with new onset CV events were on average older, more likely to be male, had higher WHR, and had higher Agatston scores (all P<0.05) (**Table 2).**

**Table 2:**
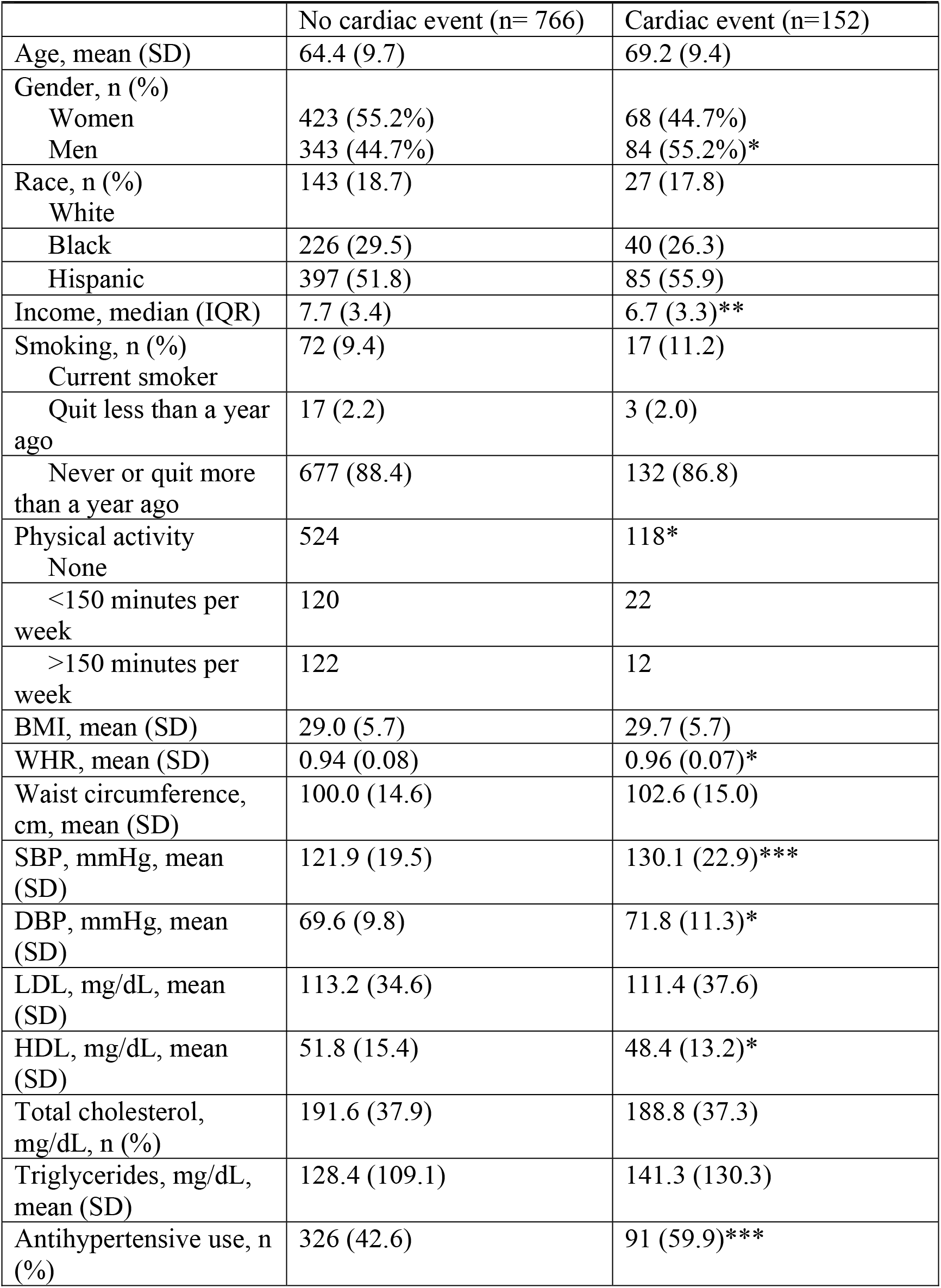
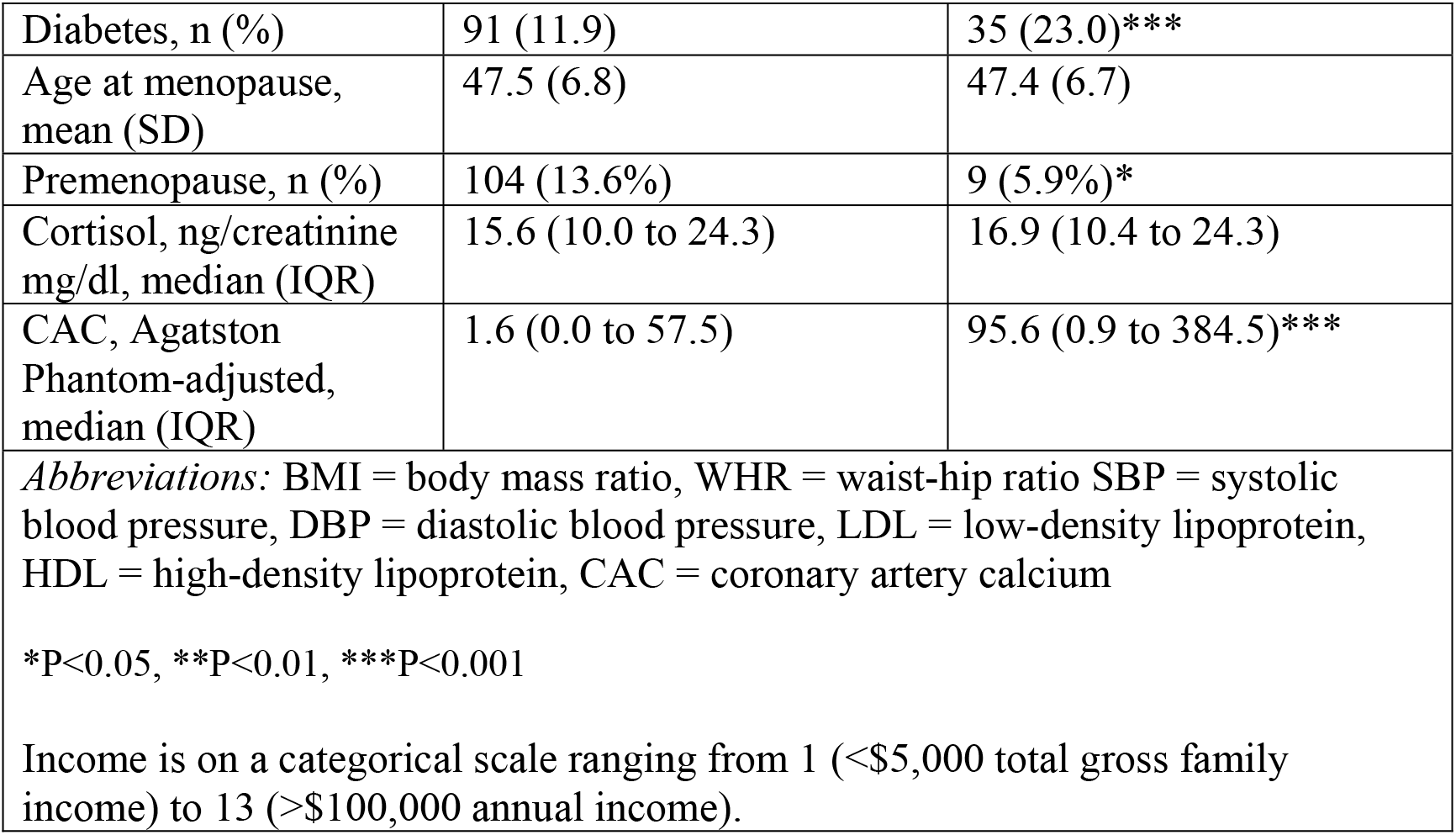
Demographics and Clinical Characteristics Stratified by New Onset Cardiovascular Events.

### Urinary cortisol, cardiovascular outcomes, and moderation by waist-hip ratio

There was no independent association between urinary cortisol and new onset CV events in the full cohort, women, or men (**Table 3**, **Table 4**). WHR had a negative moderation effect on the relationship of cortisol and new onset CV events in the whole cohort (P=0.03) and in women (P=0.03). Specifically, increased WHR was associated with a less positive association between urinary cortisol with new-onset cardiovascular events in both the full cohort and women. In women, this moderation effect had a hazard ratio of 0.67 (95% CI 0.53-0.85), indicating that the association between urinary cortisol and CV events decreases as WHR increases. On second-order moderation testing, the three-way moderation term between cortisol, gender, and WHR was statistically significant (p-value=0.03). Additional results of second-order moderation testing are described in **Supplement 1**.

**Table 3:**
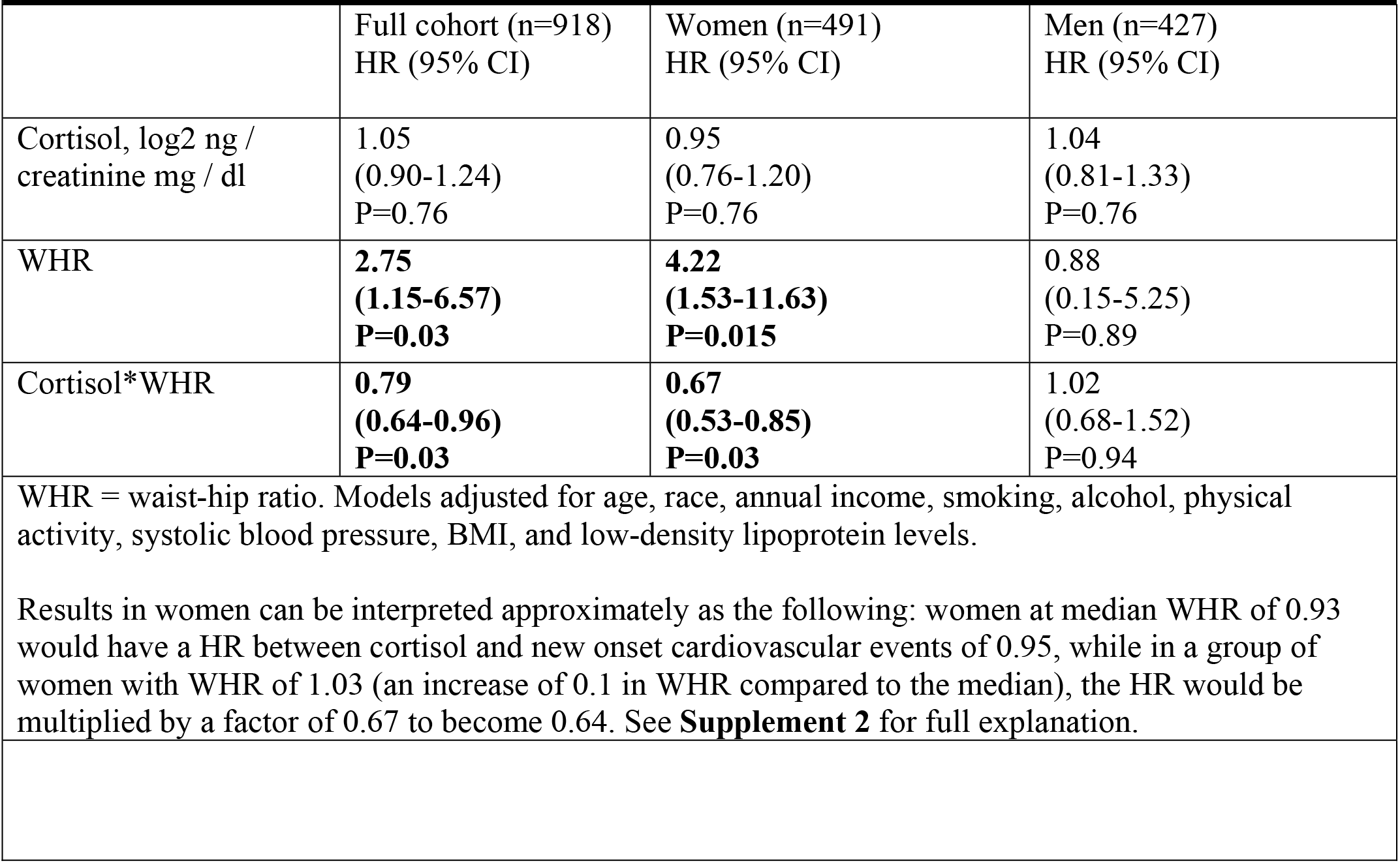
New Onset Cardiovascular events and Urinary Cortisol Moderated by Waist-Hip Ratio.

**Table 4:** Associations between New Onset Cardiovascular Events and Cortisol by Median WHR.

|  | Full cohort (n=918) |  | P for interaction | Women (n=491), |  | P for interaction | Men (n=427) |  | P for interaction |
| --- | --- | --- | --- | --- | --- | --- | --- | --- | --- |
|  | WHR≤<br>0.95,<br>N=459,<br>HR<br>(95%<br>CI) | WHR><br>0.95<br>N=459,<br>HR<br>(95%<br>CI) |  | WHR≤<br>0.93<br>N=246,<br>HR<br>(95%<br>CI) | WHR><br>0.93<br>N=245,<br>HR<br>(95%<br>CI) |  | WHR≤<br>0.98<br>N=214,<br>HR<br>(95%<br>CI) | WHR><br>0.98<br>N=213,<br>HR<br>(95%<br>CI) |  |
| Cortisol, ln ng / creatinine mg / dl | 1.23<br>(0.97-1.56)<br>P=0.09 | 0.89<br>(0.71-1.11)<br>P=0.47 | P=0.10 | 1.38<br>(0.96-1.97)<br>P=0.09 | 0.77<br>(0.56-1.07)<br>P=0.36 | **P=0.04** | 1.37<br>(0.96-1.95)<br>P=0.09 | 0.92<br>(0.67-1.27)<br>P=0.62 | P=0.23 |

All values reported are hazard ratios generated from Cox regression analyses adjusted for age, race, annual income, smoking, alcohol, physical activity, systolic blood pressure, BMI, and low-density lipoprotein levels. Cortisol was log base 2 transformed for interpretability and to meet model assumptions.

We then performed Cox multivariate regressions of the full cohort, men, and women stratified by intragroup median WHR (**Figure 1**, **Table 4**). Higher urinary cortisol tertiles were associated with increased CV events in below median WHR groups among the full cohort and trended towards significance in women and men (**Figure 1**, P=0.01, 0.06, and 0.08, respectively), but not in above median WHR groups. Specifically, in the full cohort there was a significant difference in CV events between the lowest cortisol tertile as the referent compared to either the second or third cortisol tertile (HR and 95% CI 2.56 (1.35-4.84), and 2.23 (1.17-4.28), respectively). In women, there was a significant difference between the lowest cortisol tertile and the second cortisol tertile with a borderline non-significant comparison to the third tertile (HR and 95% CI 3.72 (1.29-10.74), and 2.74 (0.95-7.90), respectively). In men, there was not a significant difference in survival between the lowest cortisol tertile and the second tertile (HR and 95% CI 1.78 (0.74-4.31)), but there was between the lowest cortisol tertile and the third cortisol tertile (2.77 (1.18-6.46)). In Cox analyses with cortisol treated as a continuous variable, higher urinary cortisol levels trended towards increased risk of new onset CV events in the below median WHR groups of the full cohort, women, and men (**Table 4**, full models, HR and 95% CI of 1.23 (0.97- 1.56), 1.38 (0.96-1.97) and 1.37 (0.96-1.95), respectively). There was significant moderation between cortisol and median WHR in women (P=0.04), but not in men or the full cohort.

**Figure 1:**
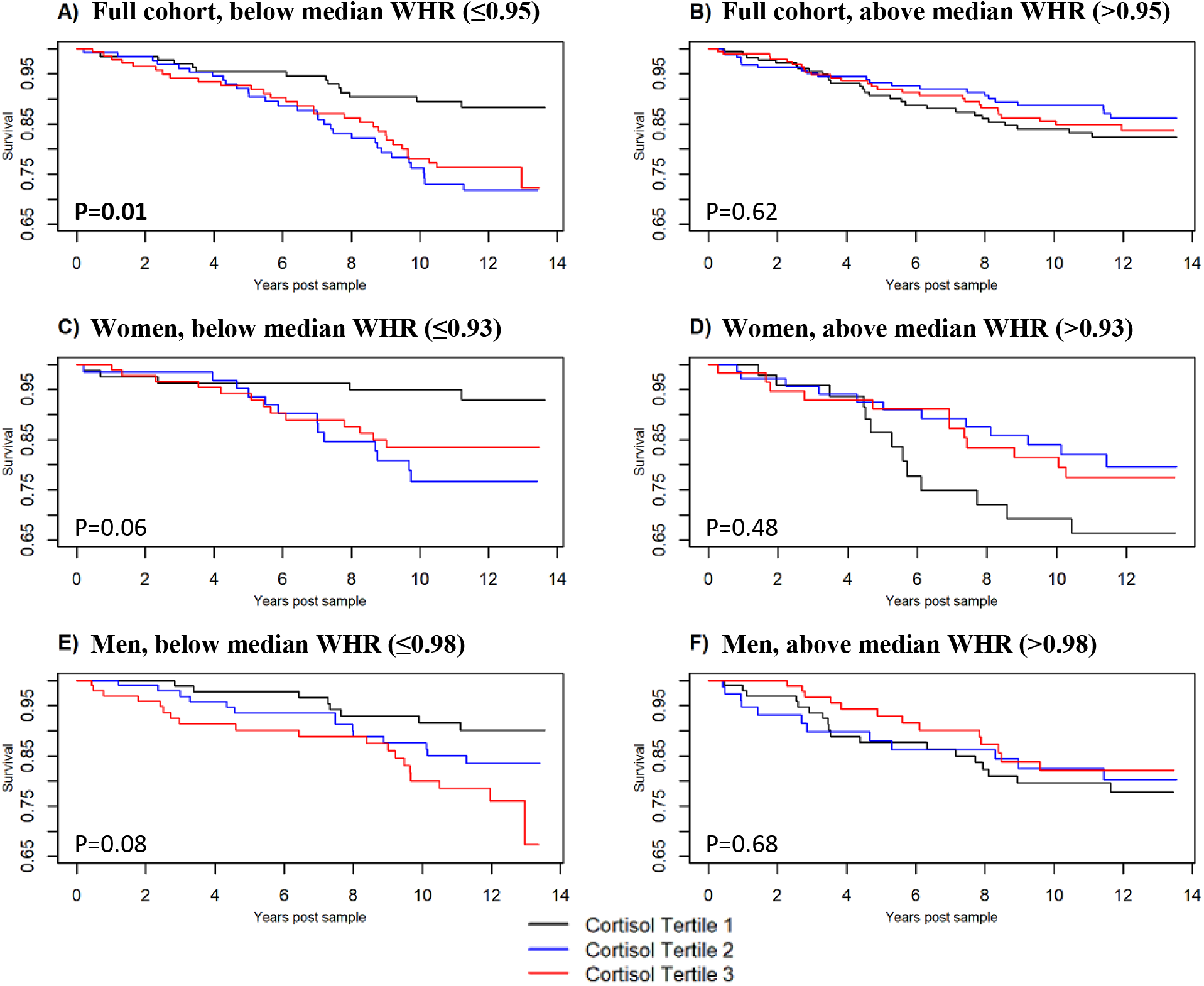
Time post urinary cortisol measurement vs. cardiovascular event-free survival by urinary cortisol tertiles, adjusted for all prespecified covariates, with columns stratified by median waist-hip ratios. P-values shown are overall significance of cortisol tertile term using likelihood ratio test. Bolding and asterisk indicate P≤0.05. HR and 95% CI for tertiles 2 and tertile 3 compared to tertile 1 as reference are: **A)** 2.56 (1.35-4.84), P=0.006; 2.34 (1.17-4.28), P=0.02, **B)** 0.76 (0.43-1.33), P=0.66; 0.89 (0.52-1.58), P=0.68, **C)** 3.72 (1.29-10.74), P=0.04; 2.74 (0.95-7.90), P=0.06, D) 0.61 (0.74-4.31), P=0.36; 0.68 (0.30-1.53), P=0.36, **E)** 1.78 (0.74-4.31), P=0.20; 2.77 (1.18-6.46), P=0.04, **F)** 0.97 (0.49-2.12), P=0.94; 0.73 (0.35-1.53), P=0.82.

Relative excess risk due to interaction (RERI), a measure of additive interaction, was calculated for all significant WHR moderation relationships in Table 3 and Table 4 and was less than zero in all comparisons. We also investigated whether BMI or waist circumference had similar moderation effects with cortisol and new onset cardiovascular events, but found no significant effects (**Supplemental Tables 1-2**).

### Urinary cortisol and change in CAC

Finally, we examined whether urinary cortisol is associated with change in CAC, and whether this change was influenced through moderation by WHR. Average time between CAC measured during MESA Stress examinations (third or fourth time point) to CAC measured at MESA fifth timepoint averaged 5.65 years, SD 0.81 years. There were 447 patients with complete CAC data. Higher urinary cortisol was associated with increased change in CAC score in the full cohort and women in the Tobit regression models (all P<0.01) but was not significant in men (P=0.21, **Table 5**). The highest association between cortisol and CAC change was in women, with an annualized relative change in CAC of 1.60 (95% CI 1.19-2.15) per 1 unit increase in ln(cortisol). There was no moderation of WHR on urinary cortisol and change in CAC in the full cohort, women, or men. Second-order moderation testing was also performed with no moderation effects. Results of this analysis are included in **Supplement 1**.

**Table 5:** Annual Change in Coronary Artery Calcium and Urinary Cortisol.

|  | Full cohort (n=447)<br>Relative change (95% CI) | Women (n=245)<br>Relative change (95% CI) | Men (n=202)<br>Relative change (95% CI) |
| --- | --- | --- | --- |
| Cortisol, log2 ng / creatinine mg / dl | **1.40**<br>**(1.14-1.72)**<br>**P=0.003** | **1.60**<br>**(1.19-2.15)**<br>**P=0.003** | 1.18<br>(0.90-1.56)<br>P=0.21 |
| WHR | 2.39<br>(0.76-7.57)<br>P=0.21 | 0.91<br>(0.20-4.13)<br>P=0.86 | 7.30<br>(0.86-61.66)<br>P=0.21 |
| Cortisol*WHR | 0.97<br>(0.74-1.27)<br>P=0.82 | 1.21<br>(0.84-1.74)<br>P=0.45 | 0.72<br>(0.45-1.16)<br>P=0.45 |

WHR = waist-hip ratio. Models include age, race, annual income, smoking, alcohol, physical activity, systolic blood pressure, BMI, and low-density lipoprotein levels. WHR was multiplied by 10 for interpretability and centered around the mean to meet model assumptions.
Results in women can be interpreted as, for every doubling in cortisol, there is expected to be a 60% increase in CAC each year. Applying this to our cohort's values for cortisol and CAC, in a one-year period women at the highest quartile of cortisol would be expected to have a median increase in CAC of about 1.12 compared to 0.6 Hounsfield units in women at the lowest quartile of cortisol. See **Supplement 3** for more detailed explanation.

## Discussion

This study analyzed a prospectively followed multi-ethnic cohort to investigate the relationship between a biological marker of stress, subclinical atherosclerosis, and new onset cardiovascular events in men and women. We also investigated whether these relationships varied by WHR, a measure of central obesity. The key findings of the present study are 1) significantly higher rates of cardiovascular events in groups with higher tertiles of urinary cortisol regardless of gender, but only among subgroups with below median WHRs, 2) a significant association between urinary cortisol and increased change in CAC in women but not men, with no effect of WHR.

Prior studies have linked urinary cortisol to cardiovascular outcomes: one study using 24-hour urine cortisol measurements that included 861 patients over age 65 and found an association with cardiovascular death, as well as a study using MESA data that included 412 patients without hypertension and identified an association with incident CVD.^9,10^ The present study found higher rates of cardiovascular events with higher urinary cortisol only in groups with lower WHR. This could be explained by a number of methodologic differences between our study and those prior studies, particularly that our study included both middle age and older adults, required that patients had no prior history of cardiovascular events, and included patients with known hypertension. Moreover, our study had a higher average BMI (29.1) compared to those two prior studies (27.5 and 28.3, respectively).

The finding that there was an association between urinary cortisol and new onset CV events only in those with low WHR has to our knowledge not been previously described. Several potential explanations may account for this finding. First, it may be that at higher WHR, the relative contribution of other cardiovascular risk factors (atherogenic lipid profiles, inflammatory adipokines, insulin resistance, etc.) outweighs any increased risk from higher levels of cortisol, masking a true underlying association between urinary cortisol and cardiovascular outcomes. Second, the increased estrogen production from greater amounts of visceral fat may moderate the relationship between urinary cortisol and CV events given estrogen’s known beneficial vascular effects,^44^ and may underlie the significant moderation effect between WHR and urinary cortisol seen only in women. Third, increased central adiposity may lead to a state of cortisol resistance. Catecholamine resistance is a known phenomenon, where in case studies of patients with pheochromocytomas higher levels of adiposity led to less hypertensive response to catecholamines.^45^ Obese individuals have also been shown to have decreased glucocorticoid feedback and unique cortisol secretion patterns, indicating a process similar to catecholamine resistance may occur with cortisol.^46,47^ There are also molecular explanations for cortisol and catecholamine resistance, including that visceral adipose tissue is high in glucocorticoid receptors and catecholamine receptors and therefore may serve as a sink for catecholamines and cortisol, preventing them from exerting their vascular or systemic effects.^21,22^ Finally, the finding may be influenced in part by statistical considerations, although we took steps to negate these effects. One statistical phenomenon is biased negative multiplicative moderation, in which when there is an outcome that is more common in those exposed to a predictor, such as WHR, the moderation effect tends to be negative. However, our RERI values for additive interaction were also consistently negative (less than zero), decreasing the likelihood that the findings were due to purely statistical phenomenon.

Although there was an association between high urinary cortisol tertile and increased CV events at below median WHR, we found a significant moderation of WHR on the relationship between cortisol and CV events only in women. Only women had significant moderation effects on both continuous and stratified analyses (see **Tables 3** and **4**). Such differences by sex may be explained by the well-established variation in adipose tissue distribution, adipose cellular characteristics, and growing clinical evidence of different health implications of adiposity in women compared men. Women are generally less likely than men to have visceral adiposity, and the increase in visceral adiposity in women is strongly linked to menopause.^18,48^ Women have also been shown to have increased adipocyte hyperplasia compared to adipocyte hypertrophy in men, and women are known to have more cardiovascular protective brown adipose tissue deposits.^18^ Increased estrogen from increased adiposity may also be more cardioprotective in women compared to men, especially since increased obesity in men is associated with lower testosterone which has been linked with CVD in men.^49^ It is important to be clear that our findings do not support the notion that increased visceral fat is cardioprotective in women; in fact, higher WHR was associated with increased CV events (P=0.015). Furthermore, WHR is an estimate rather than a direct measure of visceral fat. Rather, our findings only suggest that higher central adiposity may particularly moderate the effects of cortisol on the cardiovascular system in women.

Notably, we did find an association between increasing urinary cortisol and a higher change in CAC score in the full cohort and in women, but with no moderation by WHR in any group. Our findings linking increasing cortisol and CAC build on two previous studies that also used MESA Stress 1 data. The first demonstrated a link between salivary cortisol variation and CAC as well as ankle brachial index.^50^ More recently, we reported in a cross-sectional paper that higher urinary cortisol corresponded with greater likelihood of a CAC score>0 and higher CAC scores.^11^ Here, we show that urinary cortisol is also associated with increased change in CAC, suggesting greater rate of coronary plaque growth in individuals with higher levels of urinary cortisol. Since CAC is a robust indicator of overall atherosclerotic burden and tracks well with risk of coronary events,^51^ this finding provides evidence for a potential mechanism linking urinary cortisol levels and cardiovascular risk. There are many potential explanations underlying the increased change in CAC with increased cortisol. Cortisol increases atherogenic lipid profiles, insulin resistance, hypertension, and vascular resistance and inflammation.^7,52^ The lack of a moderation effect between WHR and cortisol with change in CAC may suggest that the moderation effect between cortisol and WHR with CV events occurs via a mechanism that is independent of coronary artery calcium accumulation.

Finally, our findings in women of an association between urinary cortisol and CAC, and of moderation by WHR on the relationship between cortisol with new onset CV events, support previous work showing that stress may have more profound CV health implications for women.^12–16^ In particular, our findings align with recent studies that demonstrated greater mental stress induced ischemia in women, especially younger women, compared to older women or men.^12–14^ Those prior studies propose a potential explanation that there is increased proclivity to microcirculatory abnormalities in women. Increased cortisol in stress potentiates the vasoconstrictor effects of catecholamines, and so may cause microvascular coronary vasoconstriction in younger women. Our findings may provide one explanation as to why the association in those prior studies was primarily identified in younger women, since younger women likely have lower visceral adiposity and therefore may experience greater effects from increased stress hormones.

We acknowledge important limitations of this study. The MESA stress study used a 12-hour overnight urine collection instead of a 24-hour urine collection, so the urinary cortisol levels may be influenced in part by sleep quality or features other than overall daily stress exposure. Another notable limitation is that urinary cortisol was measured and collected at one time point, which may limit accuracy in reflecting patient’s long-term stress levels. This also prevents analysis of trends or changes in cortisol levels, and prevents the current study from determining if those changes influence cardiovascular events or change in CAC levels. Psychological surveys of stress were also not used to corroborate findings in this study. There may also have been information bias in the measurement of waist-to-hip ratio, although this should be minimized since the measurements were performed by trained professionals in a standardized manner. The sample size of this study was relatively small since we only included MESA participants in the Stress Ancillary Study, and so conducting dose-response analyses was not possible due to limited power. Finally, our results can only identify associations rather than causations. All findings should accordingly be interpreted as exploratory and hypothesis generating.

Our findings demonstrate a significantly increased risk of CV events at higher tertiles of cortisol in below median WHR individuals regardless of gender, and a trend towards association with CV events in below median WHR groups when cortisol was treated as a continuous variable. In women, we also found an association between urinary cortisol and increased risk of subclinical atherosclerosis as measured by change in CAC. Taken together, these findings suggest stress increases atherosclerotic burden in women and that it may increase the risk of cardiovascular events in individuals with low central adiposity regardless of sex. Interestingly, central adiposity seemed to play a significant role in moderating the effects of stress on CV outcomes in women. These somewhat counterintuitive findings raise important questions about the complex interplay between sex, body composition, stress, and CVD and requires further characterization in both clinical and mechanistic studies.

## Data Availability

Data available upon request from approved MESA investigator.

## Acknowledgements

The authors thank the other investigators, the staff, and the participants of the MESA study for their valuable contributions. A full list of participating MESA investigators and institutions can be found at http://www.mesa-nhlbi.org.

## Supplemental Material

Supplement Methods, 1-3

Tables S1–S2

## References

1. Yusuf PS, Hawken S, Ôunpuu S, et al. Effect of potentially modifiable risk factors associated with myocardial infarction in 52 countries (the INTERHEART study): Case-control study. Lancet. 2004;364(9438):937–952. doi:10.1016/S0140-6736(04)17018-9

2. Ornish D, Brown SE, Billings JH, et al. Can lifestyle changes reverse coronary heart disease?. The Lifestyle Heart Trial. Lancet. 1990;336(8708):129–133. doi:10.1016/0140-6736(90)91656-U

3. Kopf S, Oikonomou D, Hartmann M, et al. Effects of stress reduction on cardiovascular risk factors in type 2 diabetes patients with early kidney disease - Results of a randomized controlled trial (HEIDIS). Exp Clin Endocrinol Diabetes. 2014;122(6):341–349. doi:10.1055/s-0034-1372583

4. Chandola T, Britton A, Brunner E, et al. Work stress and coronary heart disease: What are the mechanisms? Eur Heart J. 2008;29(5):640–648. doi:10.1093/eurheartj/ehm584

5. C C-D, AV DR, T S, S S, S S, S T. Associations of socioeconomic and psychosocial factors with urinary measures of cortisol and catecholamines in the Multi-Ethnic Study of Atherosclerosis (MESA). Psychoneuroendocrinology. 2014;41. doi:10.1016/J.PSYNEUEN.2013.12.013

6. Lupien SJ, King S, Meaney MJ, McEwen BS. Can poverty get under your skin? Basal cortisol levels and cognitive function in children from low and high socioeconomic status. Dev Psychopathol. 2001;13(3):653–676. doi:10.1017/S0954579401003133

7. Pickering T. Cardiovascular pathways: Socioeconomic status and stress effects on hypertension and cardiovascular function. In: Annals of the New York Academy of Sciences. Vol 896. New York Academy of Sciences; 1999:262–277. doi:10.1111/j.1749-6632.1999.tb08121.x

8. Steptoe A, Kivimäki M. Stress and cardiovascular disease. Nat Rev Cardiol. 2012;9(6):360-370. doi:10.1038/nrcardio.2012.45

9. Vogelzangs N, Beekman ATF, Milaneschi Y, Bandinelli S, Ferrucci L, Penninx BWJH. Urinary cortisol and six-year risk of all-cause and cardiovascular mortality. J Clin Endocrinol Metab. 2010;95(11):4959–4964. doi:10.1210/jc.2010-0192

10. Inoue K, Horwich T, Bhatnagar R, et al. Urinary Stress Hormones, Hypertension, and Cardiovascular Events: The Multi-Ethnic Study of Atherosclerosis. Hypertension. 2021;78:1640–1647. doi:10.1161/HYPERTENSIONAHA.121.17618

11. Zipursky RT, Press MC, Srikanthan P, et al. Relation of Stress Hormones (Urinary Catecholamines/Cortisol) to Coronary Artery Calcium in Men Versus Women (from the Multi-Ethnic Study of Atherosclerosis [MESA]). Am J Cardiol. 2017;119(12):1963–1971. doi:10.1016/j.amjcard.2017.03.025

12. Vaccarino V, Wilmot K, Mheid I Al, et al. Sex differences in mental stress-induced myocardial ischemia in patients with coronary heart disease. J Am Heart Assoc. 2016;5(9). doi:10.1161/JAHA.116.003630

13. Vaccarino V, Sullivan S, Hammadah M, et al. Mental Stress–Induced-Myocardial Ischemia in Young Patients With Recent Myocardial Infarction. Circulation. 2018;137(8):794–805. doi:10.1161/CIRCULATIONAHA.117.030849

14. Samad Z, Boyle S, Ersboll M, et al. Sex Differences in Platelet Reactivity and Cardiovascular and Psychological Response to Mental Stress in Patients With Stable Ischemic Heart Disease: Insights From the REMIT (Responses of Mental Stress–Induced Myocardial Ischemia to Escitalopram) Study. J Am Coll Cardiol. 2014;64(16):1669. doi:10.1016/J.JACC.2014.04.087

15. Saab PG, Matthews KA, Stoney CM, McDonald RH. Premenopausal and Postmenopausal Women Differ in their Cardiovascular and Neuroendocrine Responses to Behavioral Stressors. Psychophysiology. 1989;26(3):270–280. doi:10.1111/j.1469-8986.1989.tb01917.x

16. Fonseca MIH, Da Silva IT, Ferreira SRG. Impact of menopause and diabetes on atherogenic lipid profile: is it worth to analyse lipoprotein subfractions to assess cardiovascular risk in women? Diabetol Metab Syndr. 2017;9(1):22. doi:10.1186/S13098-017-0221-5

17. Abildgaard J, Ploug T, Al-Saoudi E, et al. Changes in abdominal subcutaneous adipose tissue phenotype following menopause is associated with increased visceral fat mass. Sci Reports 2021 111. 2021;11(1):1–12. doi:10.1038/S41598-021-94189-2

18. Palmer BF, Clegg DJ. The sexual dimorphism of obesity. Mol Cell Endocrinol. 2015;0:113. doi:10.1016/J.MCE.2014.11.029

19. Després JP. Health consequences of visceral obesity. Ann Med. 2001;33(8):534–541. doi:10.3109/07853890108995963

20. Abraham TM, Pedley A, Massaro JM, Hoffmann U, Fox CS. Association between visceral and subcutaneous adipose depots and incident cardiovascular disease risk factors. Circulation. 2015;132(17):1639–1647. doi:10.1161/CIRCULATIONAHA.114.015000

21. Björntorp P. Hormonal control of regional fat distribution. Hum Reprod. 1997;12 Suppl 1:21–25. doi:10.1093/humrep/12.suppl_1.21

22. Ibrahim MM. Subcutaneous and visceral adipose tissue: structural and functional differences. Obes Rev. 2010;11(1):11–18. doi:10.1111/J.1467-789X.2009.00623.X

23. Brunner EJ, Hemingway H, Walker BR, et al. Adrenocortical, autonomic, and inflammatory causes of the metabolic syndrome: nested case-control study. Circulation. 2002;106(21):2659–2665. doi:10.1161/01.CIR.0000038364.26310.BD

24. ES E, B M, T S, et al. Stress and body shape: stress-induced cortisol secretion is consistently greater among women with central fat. Psychosom Med. 2000;62(5):623–632. doi:10.1097/00006842-200009000-00005

25. Pasquali R, Vicennati V. Activity of the hypothalamic-pituitary-adrenal axis in different obesity phenotypes. Int J Obes Relat Metab Disord. 2000;24 Suppl 2:S47–S49. doi:10.1038/sj.ijo.0801277

26. Ward AMV, Fall CHD, Stein CE, et al. Cortisol and the metabolic syndrome in South Asians. Clin Endocrinol (Oxf). 2003;58(4):500. doi:10.1046/J.1365-2265.2003.01750.X

27. Korbonits M, Trainer PJ, Nelson ML, et al. Differential stimulation of corticol and dehydropiandrosterone levels by food in obese and normal subjects: relation to body fat distribution. Clin Endocrinol (Oxf). 1996;45(6):699–706. doi:10.1046/J.1365-2265.1996.8550865.X

28. Pasquali R, Anconetani B, Chattat R, et al. Hypothalamic-pituitary-adrenal axis activity and its relationship to the autonomic nervous system in women with visceral and subcutaneous obesity: Effects of the corticotropin-releasing factor/arginine-vasopressin test and of stress. Metab - Clin Exp. 1996;45(3):351–356. doi:10.1016/S0026-0495(96)90290-5

29. Pasquali R, Ambrosi B, Armanini D, et al. Cortisol and ACTH Response to Oral Dexamethasone in Obesity and Effects of Sex, Body Fat Distribution, and Dexamethasone Concentrations: A Dose-Response Study. J Clin Endocrinol Metab. 2002;87(1):166–175. doi:10.1210/JCEM.87.1.8158

30. Misra M, Bredella MA, Tsai P, Mendes N, Miller KK, Klibanski A. Lower growth hormone and higher cortisol are associated with greater visceral adiposity, intramyocellular lipids, and insulin resistance in overweight girls. Am J Physiol - Endocrinol Metab. 2008;295(2):E385. doi:10.1152/AJPENDO.00052.2008

31. Mårin P, Darin N, Amemiya T, Andersson B, Jern S, Björntorp P. Cortisol secretion in relation to body fat distribution in obese premenopausal women. Metabolism. 1992;41(8):882–886. doi:10.1016/0026-0495(92)90171-6

32. Pasquali R, Cantobelli S, Casimirri F, et al. The hypothalamic-pituitary-adrenal axis in obese women with different patterns of body fat distribution. J Clin Endocrinol Metab. 1993;77(2):341–346. doi:10.1210/JCEM.77.2.8393881

33. Boscaro M, Giacchetti G, Ronconi V. Visceral adipose tissue: emerging role of gluco-and mineralocorticoid hormones in the setting of cardiometabolic alterations. Ann N Y Acad Sci. 2012;1264(1):87. doi:10.1111/J.1749-6632.2012.06597.X

34. Bild DE, Bluemke DA, Burke GL, et al. Multi-Ethnic Study of Atherosclerosis: Objectives and design. Am J Epidemiol. 2002;156(9):871–881. doi:10.1093/aje/kwf113

35. Castro-Diehl C, Roux AVD, Seeman T, Shea S, Shrager S, Tadros S. Associations of socioeconomic and psychosocial factors with urinary measures of cortisol and catecholamines in the Multi-Ethnic Study of Atherosclerosis (MESA). Psychoneuroendocrinology. 2014;41:132–141. doi:10.1016/j.psyneuen.2013.12.013

36. Seeman TE, Crimmins E, Huang MH, et al. Cumulative biological risk and socio-economic differences in mortality: MacArthur studies of successful aging. Soc Sci Med. 2004;58(10):1985–1997. doi:10.1016/S0277-9536(03)00402-7

37. Ambale-Venkatesh B, Yang X, Wu CO, et al. Cardiovascular Event Prediction by Machine Learning: The Multi-Ethnic Study of Atherosclerosis. Circ Res. 2017;121(9):1092. doi:10.1161/CIRCRESAHA.117.311312

38. Inoue K, Goldwater D, Allison M, Seeman T, Kestenbaum BR, Watson KE. Serum Aldosterone Concentration, Blood Pressure, and Coronary Artery Calcium: The Multi-Ethnic Study of Atherosclerosis. Hypertension. 2020;76(1):113–120. doi:10.1161/HYPERTENSIONAHA.120.15006

39. Detrano R, Guerci AD, Carr JJ, et al. Coronary Calcium as a Predictor of Coronary Events in Four Racial or Ethnic Groups. http://dx.doi.org/101056/NEJMoa072100. 2009;358(13):1336–1345. doi:10.1056/NEJMOA072100

40. Agatston AS, Janowitz WR, Hildner FJ, Zusmer NR, Viamonte M, Detrano R. Quantification of coronary artery calcium using ultrafast computed tomography. J Am Coll Cardiol. 1990;15(4):827–832. doi:10.1016/0735-1097(90)90282-T

41. Alluri K, Joshi PH, Henry TS, Blumenthal RS, Nasir K, Blaha MJ. Scoring of coronary artery calcium scans: History, assumptions, current limitations, and future directions. Atherosclerosis. 2015;239(1):109–117. doi:10.1016/J.ATHEROSCLEROSIS.2014.12.040

42. Vaidya D, Dobs A, Gapstur SM, et al. Association of Baseline Sex Hormone Levels with Baseline and Longitudinal Changes in Waist-to-Hip Ratio : Multi-Ethnic Study of Atherosclerosis. Int J Obes (Lond). 2012;36(12):1578. doi:10.1038/IJO.2012.3

43. See R, Abdullah SM, McGuire DK, et al. The association of differing measures of overweight and obesity with prevalent atherosclerosis: the Dallas Heart Study. J Am Coll Cardiol. 2007;50(8):752–759. doi:10.1016/J.JACC.2007.04.066

44. Murphy E. Estrogen signaling and cardiovascular disease. Circ Res. 2011;109(6):687-696. doi:10.1161/CIRCRESAHA.110.236687/FORMAT/EPUB

45. LEE RE, ROUSSEAU P. Pheochromocytoma and Obesity. J Clin Endocrinol Metab. 1967;27(7):1050-1052. doi:10.1210/jcem-27-7-1050

46. Jessop DS, Dallman MF, Fleming D, Lightman SL. Resistance to glucocorticoid feedback in obesity. J Clin Endocrinol Metab. 2001;86(9):4109–4114. doi:10.1210/JCEM.86.9.7826

47. Björntorp P, Rosmond R. Obesity and cortisol. Nutrition. 2000;16(10):924-936. doi:10.1016/S0899-9007(00)00422-6

48. Manolopoulos KN, Karpe F, Frayn KN. Gluteofemoral body fat as a determinant of metabolic health. Int J Obes 2010 346. 2010;34(6):949–959. doi:10.1038/ijo.2009.286

49. Grossmann M, Tang Fui M, Dupuis P. Lowered testosterone in male obesity: Mechanisms, morbidity and management. Asian J Androl. 2014;16(2):223. doi:10.4103/1008-682X.122365

50. Hajat A, Diez-Roux A V., Sánchez BN, et al. Examining the association between salivary cortisol levels and subclinical measures of atherosclerosis: The Multi-Ethnic Study of Atherosclerosis. Psychoneuroendocrinology. 2013;38(7):1036–1046. doi:10.1016/j.psyneuen.2012.10.007

51. Kondos GT, Hoff JA, Sevrukov A, et al. Electron-Beam Tomography Coronary Artery Calcium and Cardiac Events. Circulation. 2003;107(20):2571–2576. doi:10.1161/01.CIR.0000068341.61180.55

52. Steptoe A, Kivimäki M. Stress and cardiovascular disease. Nat Rev Cardiol 2012 96. 2012;9(6):360–370. doi:10.1038/nrcardio.2012.45

